# Rejecting Perfection, Valuing Imperfection in Perinatal Care: a Mixed-Methods Study of Responsive Practice Through The Five Misfits by Beatrice Alemagna

**DOI:** 10.64898/2026.02.06.26345631

**Authors:** Claudia Ravaldi, Alfredo Vannacci

## Abstract

Picture books are used in the education of health professionals, but little is known about how the structure of a specific artwork shapes what professionals bring to reflection. We examined how perinatal professionals read Beatrice Alemagna’s *The Five Misfits*, a fable in which five imperfect characters face a self-proclaimed Perfect One, and we argue that the book did its most useful work where it leaves a character’s value unstated. Thirty-four women attending an Italian postgraduate course on the first thousand days (psychologists, midwives, nurses, pharmacists, early childhood educators and one physician) wrote forum reflections on the characters. Reflections were analysed thematically, eight dimensions of responsive care were scored on a bipolar scale (−2 to +2), and a 90-minute facilitated session used Plutchik’s wheel of emotions to explore reactions to the most rejected misfit. Participants rejected the Perfect One (90% negative feelings) as a paternalistic anti-model and valued four misfits (73–95% positive feelings) for complementary qualities, largely adopting the verdicts the book gives. The Feeble One, the only misfit who claims no strength, drew 85% negative feelings and the highest self-reflection scores. In the facilitated session 23 participants gave 59 emotional attributions, mostly of sadness and anger; through discussion they named these reactions and distinguished passivity linked to depression from other forms of withdrawal. We conclude that a picture book can bring tacit professional norms into view, above all where its narrative withholds judgement: there readers supply their own, and a facilitator can turn that judgement into an object of reflection.

## 1 Introduction

Literature, visual narrative and picture books are increasingly used in the education of health professionals to support empathy and reflective practice. Systematic reviews of narrative medicine programmes, which combine close reading, reflective writing and story-telling, report high participant satisfaction and gains in perspective-taking, professional identity and relationship-building (Milota et al. 2019; Remein et al. 2020; Xue et al. 2023; Palla et al. 2024). Studies with nursing students describe visual narratives as a stimulus to imagination and to engagement with patients’ experiences (Stone and Levett-Jones 2014; Williams et al. 2024). The same reviews note that outcomes are inconsistent and that the qualitative methods used to study these programmes are often weak (Milota et al. 2019; Morrison 2023). Less attention has gone to the artwork itself, that is, to how the structure of a particular book shapes what its readers bring to it.

Picture books hold a particular place among these resources. They combine images and words, so that readers attend at once to what the pictures show and to what the text says, and the best of them treat fundamental human experiences through image and metaphor without becoming didactic. Their apparent simplicity leaves room for interpretation, and in our earlier work perinatal professionals used that room to engage with complex caregiving concepts (Ravaldi and Vannacci 2025).

Perinatal care is a demanding setting in which to ask this question, because responsive caregiving, the capacity to understand and answer a child’s cues and needs, is associated with better socioemotional outcomes in children (Scherer et al. 2019), and the professionals who support parents in the first years of life need a comparable attunement towards families. They work under considerable emotional strain: in our own research, nearly half of the midwives working in facilities exposed to perinatal loss reported symptoms consistent with burnout (Ravaldi et al. 2022).

In a previous study with the same cohort, participants used the metaphorical language of Alemagna’s *What is a Child?* to articulate principles of nurturing care and to explore their professional identities (Ravaldi and Vannacci 2025). That book is affirming in tone and gave little access to the uncomfortable side of professional identity, whereas *The Five Misfits* (*I cinque malfatti*, 2014; English translation 2017) is built differently (Alemagna 2014, 2017). Five imperfect characters live contentedly in a crooked house until a self-proclaimed Perfect One arrives and dismisses them as worthless; the misfits answer by asserting the value of their apparent defects, and the Perfect One is left alone. The fable bears on perfectionism, paternalism and the worth of imperfection, which our interpretative work places at the centre of responsive care (Ravaldi 2024, 2025, 2026; Vannacci 2026). Metaphor can bring to light the unwritten assumptions that shape professional behaviour (Aita et al. 2003; Daley 2001), and storytelling can make relational dynamics available for examination (Gray 2009).

We argue that the book did its most useful work where it is silent. Its characters are archetypes, and for five of the six the narrative states a verdict that readers can adopt. The sixth, the Feeble One, falls asleep when the misfits are asked what they can offer, and the book never says what he is worth. Reading the forum, we found that participants followed the book where it evaluates and that, where it does not, their own professional norms came into view. We therefore asked how perinatal professionals interpreted the six characters, and how a facilitated discussion of their reactions to the Feeble One supported reflection on responsive care.

## 2 Methods

We aimed to describe how perinatal professionals interpreted the six characters of *The Five Misfits*, to compare the qualities of responsive care they attributed to each character, and to explore how a facilitated discussion of their reactions to the Feeble One supported reflection on their practice.

### 2.1 Setting and participants

We analysed forum discussions from a practice-oriented module of a postgraduate specialisation course on early childhood development and perinatal care (*I Primi Mille Giorni*, The First Thousand Days, offered by the University of Florence since 2023). This is the second of a series of studies on picture books in professional development; the first used *What is a Child?* with the same cohort (Ravaldi and Vannacci 2025). The forum opened after a live presentation of the book and a general overview of responsive care principles. We kept this introduction deliberately broad, without guidance on interpretation, so that participants could build their own connections between the book and their practice.

### 2.2 The picture book

In *I cinque malfatti* five unusual characters share a crooked house: il Bucato (the Punctured One, full of holes), il Piegato (the Folded One, covered in creases), il Molle (the Feeble One, always tired and soft), il Capovolto (the Upside-Down One, who walks on his hands) and lo Sbagliato (the Wrong One, a complete disaster). Il Perfetto (the Perfect One) arrives, criticises them and proposes solutions to ‘fix’ their defects. Four misfits reply by naming a strength: the Punctured One lets anger pass through his holes, the Folded One keeps precious memories in his creases, the Upside-Down One sees things others cannot see, and the Wrong One celebrates every small success with a party, while the Feeble One answers ‘Zzz…’. The Perfect One departs ‘like a truly perfect fool’, and the misfits remain together in their imperfect house.

### 2.3 Forum prompts and data collection

Participants answered two prompts: (1) Who is the responsive healthcare professional? What does she or he do and not do? What does she or he bring and avoid bringing into the relationship with parents and their child? (2) Indicate one misfit with whom you feel in harmony and one that repels you. Why? Initial reflections were due by a set date, and although the forum was visible from the outset, most participants posted close to the deadline, which limited conformity in these first posts; they then had one further month to elaborate their thoughts in conversation with peers.

### 2.4 Thematic analysis and scoring

We combined a qualitative thematic analysis with a quantitative scoring of the same material. The thematic analysis preserved the nuance of participants’ reflections, and the scoring allowed us to compare systematically how participants attributed professional qualities to the different characters and how those attributions varied across professional groups.

The qualitative analysis was primarily inductive, because our study of *What is a Child?* had used deductive coding guided by the WHO Nurturing Care Framework (Ravaldi and Vannacci 2025) and here we wanted to see how participants spontaneously connected the characters to their practice. An initial AI-assisted coding followed a previously described method (Ravaldi et al. 2023a; Ravaldi and Vannacci 2025), and the research team then reviewed, refined and discussed the themes through manual coding in MAXQDA 2018 (VERBI Software GmbH, Berlin, Germany). Through iterative refinement we identified eight dimensions of responsive caregiving: Patient Autonomy (respect for family self-determination versus paternalistic imposition), Relational Attunement (empathic listening and therapeutic alliance), Personalisation (care adapted to individual needs), Professional Humility (recognition of one’s limits and of teamwork), Emotional Processing (constructive management of emotions), Perspective Taking (seeing situations from unconventional angles), Initiative/Activation (proactive engagement versus passivity) and Self-Reflection (personal identification and introspection triggered by the character).

Each participant’s reflection on each character, and her description of the responsive caregiver, which served as a normative benchmark, was scored on the eight dimensions with a bipolar scale from −2 to +2. Positive values mean that the participant attributed the quality to the character in a favourable sense (for example, the character listens well), negative values that she attributed it in a problematic sense (the character fails to listen or imposes his views), and zero that the theme was absent.

To map the reception of the characters we built two composite indices. Perceived Emotional Connection (Y-axis) is the percentage of positive feelings attributed to a character minus 50, so that the scale is centred on zero. Perceived Personal Dynamism (X-axis) combines the Initiative/Activation score, weighted ×30, with a normalised measure of cognitive dynamism (Emotional Processing plus Perspective Taking).

### 2.5 Facilitated discussion on the Feeble One

The Feeble One drew the strongest negative reactions in the forum, and we devoted to him a 90-minute synchronous session. Participants imagined him both as a colleague and as a patient in their care, identified their emotional reactions on Plutchik’s wheel of emotions (Plutchik 1982) and rated the intensity of each from 1 (minimal) to 10 (overwhelming). The exercise was at once a means of data collection and a bibliotherapeutic intervention, in which participants could acknowledge and work through their negative reactions in a facilitated group. We transcribed the discussion and analysed it thematically, looking at the emotions elicited, at the two readings of the character (colleague and patient) and at the changes of view that emerged during the session.

### 2.6 Statistical analysis

Quantitative analyses were run in Stata BE 18 (Stata-Corp), with thematic scores treated as approximately interval-level summary measures, given the symmetrical scale, and reported as means and standard deviations; feelings and emotions are reported as frequencies and percentages. Differences in thematic profiles across characters were assessed with a linear mixed-effects model with random intercepts for participants, to account for repeated evaluations by the same person, and tested with a Wald χ^2^ test. Differences across characters on single dimensions, and across professional groups, were examined with Kruskal–Wallis tests. Effect sizes were computed in R (version 4.6.1) with the effectsize and performance packages and are reported as epsilon squared (ε^2^) with bootstrap 95% confidence intervals (2,000 resamples) for Kruskal–Wallis tests and as marginal R^2^ for the mixed-effects model. We report exact p values and read the results through the size and precision of the estimates. Intensity ratings from the Plutchik exercise are reported as individual values with their emotion labels.

### 2.7 Ethical considerations

The forum contributions were anonymised before analysis and no personal data were collected. Participants consented to the use of their forum posts for research purposes, as stated in the forum’s participation guidelines, and all of them are named in the Acknowledgements. Review by an ethics committee was not sought.

### 2.8 Use of artificial intelligence

ChatGPT 5.2 (OpenAI) and Claude Sonnet and Opus 4.5 (Anthropic) supported the initial coding and the thematic quantification of the forum discussions, as described above, and the translation of specific passages from Italian into English; Claude Opus 5.5 (Anthropic) supported the editing and shortening of the manuscript. The authors directed and checked every use of these tools and take sole responsibility for the analysis and the text.

## 3 Results

### 3.1 Participants and character reception

Thirty-four women took part (median age 34 years, interquartile range 29–39, range 25–71): 14 psychologists, 11 midwives, three pharmacists, three early childhood educators, two nurses and one physician. The forum threads, opened by the two authors with open-ended prompts, produced 126 posts and 19,783 words.

Most participants rejected the Perfect One and the Feeble One, with 90% and 85% negative feelings respectively, while the four other misfits were valued, with 73% to 95% positive feelings (Figure 1). Thematic profiles differed across characters in the mixed-effects model (Wald χ^2^(6)=38.47, p<0.001; marginal R^2^=0.04). Professional groups did not differ in their scores (Kruskal–Wallis χ^2^=3.82, p=0.576; ε^2^=0.004, 95% CI 0.002–0.021), and psychologists, midwives, educators and the other professionals read the characters in much the same way.

**Figure 1.**
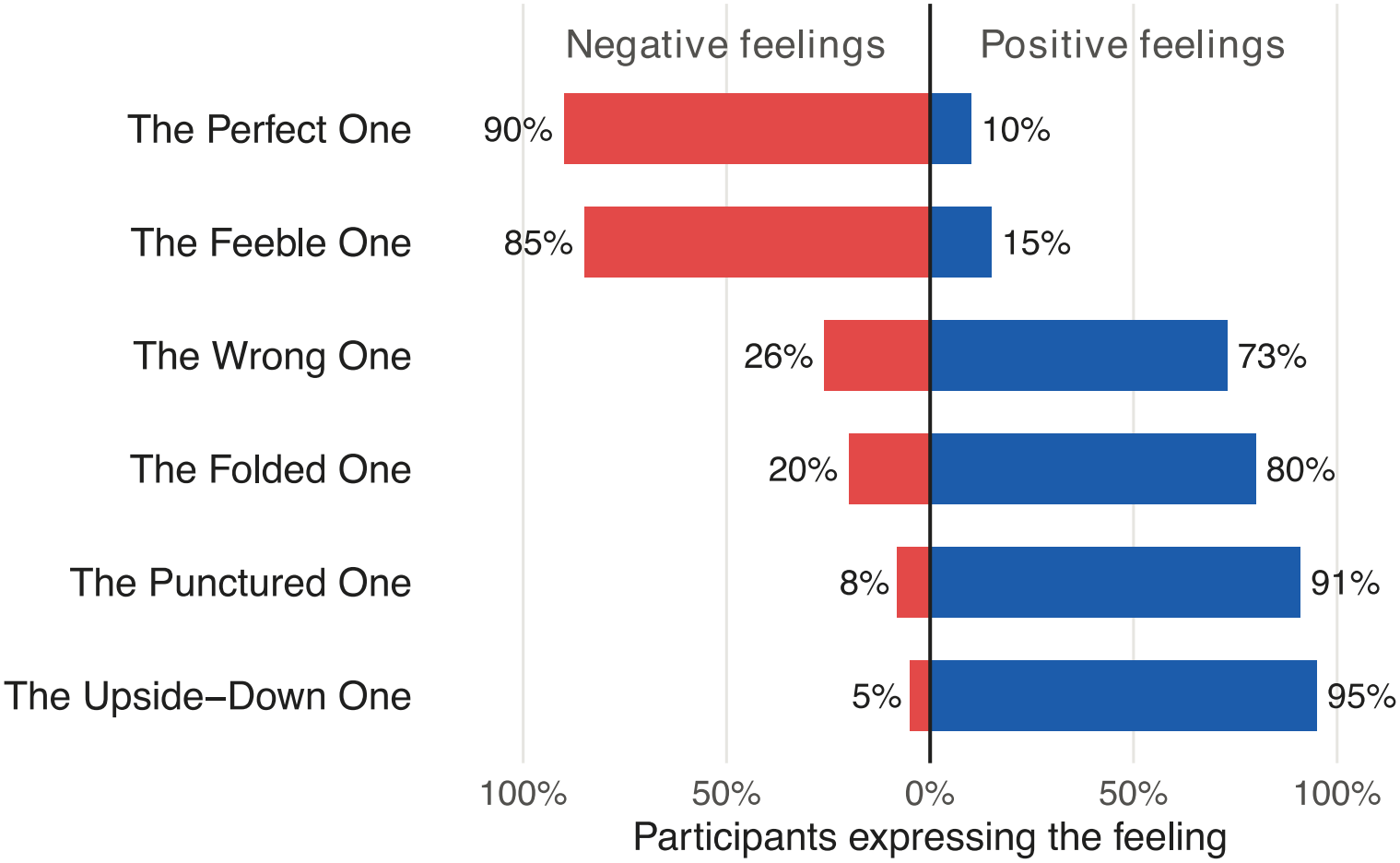
Overview of character reception: positive and negative feelings. Proportion of participants expressing positive (right) or negative (left) feelings towards each character.

### 3.2 The responsive caregiver as benchmark

Participants described the responsive caregiver through high scores on Relational Attunement (mean 1.56, SD 0.61), Personalisation (mean 1.32, SD 0.64) and Professional Humility (mean 0.88, SD 0.73), and through an explicit critique of paternalism (Patient Autonomy mean −1.21, SD 0.77, where the negative value records that criticism and not the absence of the theme). Participants placed the responsive professional alongside families, listening without imposing predetermined solutions and aware of the limits of her own expertise, as a midwife wrote:

The responsive carer is one who knows how to stand beside the person, or the family if we are talking about the first thousand days, ‘without placing themselves above.’ Using their specific competencies to the fullest, they know how to establish an authentic and respectful professional relationship with those they assist. At the same time, they recognise their own limits and the importance of networking.

Table 1 reports the profiles of the benchmark and of the six characters, and in what follows the space we give to each character below follows the number and density of the responses it generated: the Feeble One drew the strongest reactions and the most elaborate reflections, while the Folded One, for example, was mentioned more sparingly.

**Table 1.** Thematic scores by character and statistical comparisons.

| **Character** | **Patient Autonomy** | **Relational Attunement** | **Personal-isation** | **Professional Humility** | **Emotional Processing** | **Perspective Taking** | **Initiative / Activation** | **Self-Reflection** |
|---|---|---|---|---|---|---|---|---|
| Responsive Caregiver | −1.21 (0.77) | 1.56 (0.61) | 1.32 (0.64) | 0.88 (0.73) | 0.41 (0.65) | 0.47 (0.66) | 0.71 (0.72) | 0.26 (0.56) |
| Perfect One | −1.62 (0.51) | 0.62 (0.77) | 0.62 (0.65) | 0.46 (0.66) | 0.31 (0.63) | 0.69 (0.63) | --- | 0.54 (0.78) |
| Punctured One | −0.12 (0.33) | 0.25 (0.45) | 0.06 (0.25) | 0.31 (0.48) | 1.31 (0.79) | 0.25 (0.45) | --- | 0.62 (0.62) |
| Folded One | --- | --- | 0.25 (0.45) | 0.25 (0.45) | 0.08 (0.29) | 1.33 (0.65) | --- | 0.25 (0.45) |
| Feeble One | --- | 0.12 (0.33) | 0.04 (0.20) | 0.25 (0.44) | 0.25 (0.53) | 0.29 (0.55) | −1.71 (0.46) | 1.00 (0.72) |
| Upside Down One | --- | 0.33 (0.49) | 0.17 (0.39) | 0.17 (0.39) | 0.08 (0.29) | 1.92 (0.29) | --- | 0.42 (0.51) |
| Wrong One | 0.07 (0.26) | --- | 0.07 (0.26) | 0.07 (0.26) | 1.13 (0.74) | 0.27 (0.46) | −0.20 (0.56) | 0.53 (0.64) |

Values are mean (SD). Scores range from −2 to +2. Null values “0.00 (0.00)” are reported as “---" for readability. Differences between characters were tested with a mixed-effects model with random intercepts for participants (Wald χ<sup>2</sup>(6)=38.47, p<0.001; marginal R<sup>2</sup>=0.04); differences between thematic dimensions (χ<sup>2</sup>=245.59, p<0.001; ε<sup>2</sup>=0.24, 95% CI 0.22–0.28) and across professional groups (χ<sup>2</sup>=3.82, p=0.576; ε<sup>2</sup>=0.004, 95% CI 0.002–0.021) with Kruskal–Wallis tests.

### 3.3. Mapping character reception

On the two composite dimensions the rejected and the valued characters fall apart (Figure 2), with the Feeble One and the Perfect One in the lower-left quadrant. The Feeble One has the most negative Personal Dynamism, in line with his Initiative/Activation score (mean −1.71, SD 0.46), while the Perfect One is closer to neutral on dynamism and lowest on Emotional Connection. The four valued misfits sit in the upper half of the map, near the centre of the dynamism axis. The Upside-Down One occupies the most favourable position, with the highest Perspective Taking score (mean 1.92, SD 0.29) and the highest share of positive feelings (95%); the Punctured One, with the highest Emotional Processing score (mean 1.31, SD 0.79), also lies on the favourable side of both dimensions.

**Figure 2.**
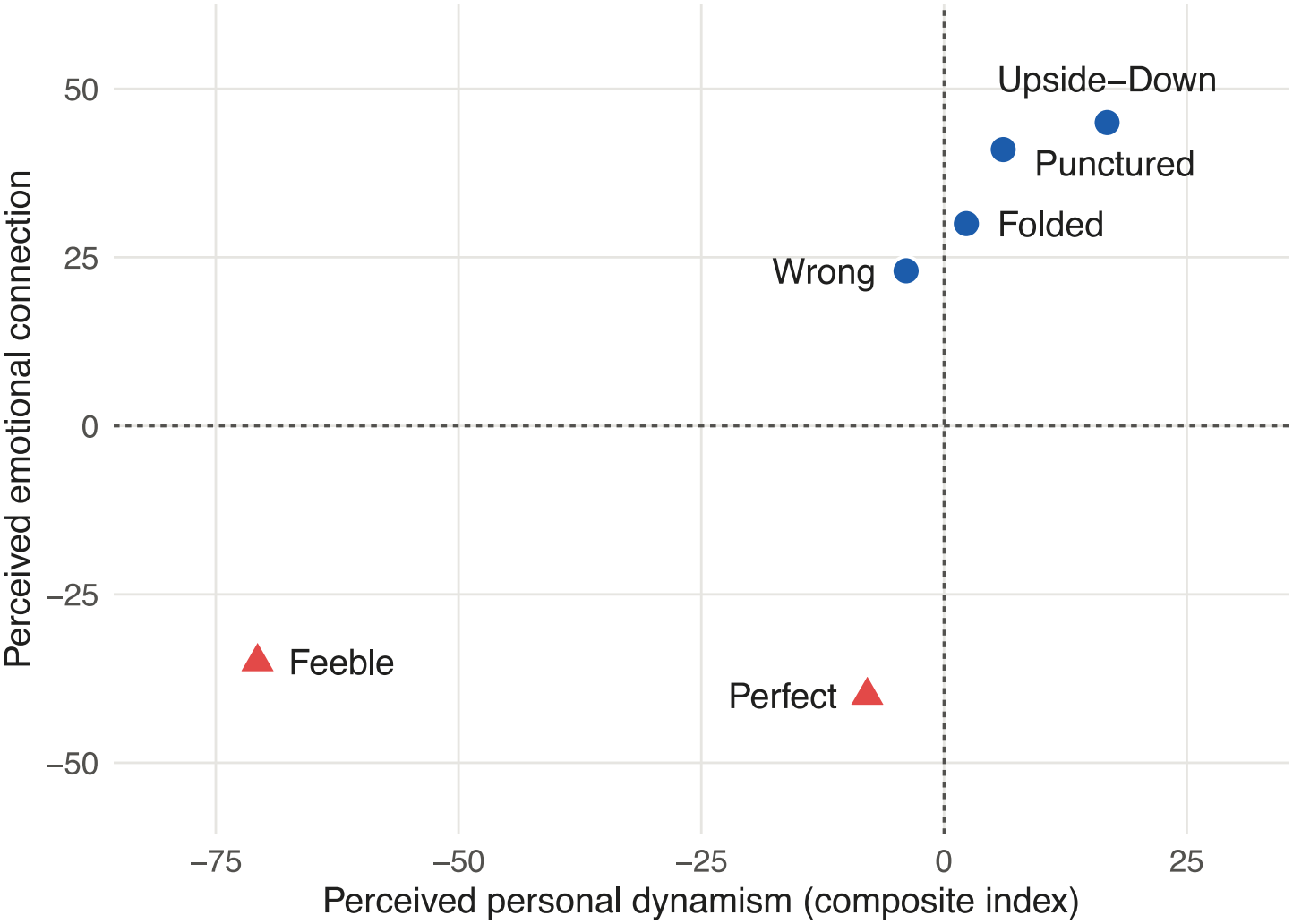
Mapping the misfits on two dimensions of responsive caregiving qualities. The Y-axis (Perceived Emotional Connection) derives from the proportion of positive versus negative feelings attributed to each character; the X-axis (Perceived Personal Dynamism) combines Initiative/Activation scores with Emotional Processing and Perspective Taking. Rejected characters (Perfect, Feeble) occupy the lower-left quadrant; valued characters lie in the upper half.

### 3.4 The Perfect One and the valued misfits

The Perfect One had the most negative Patient Autonomy score (mean −1.62, SD 0.51), and the characters differed on this dimension more than on any other except Initiative/Activation (Kruskal–Wallis χ^2^=90.69, p<0.001; ε^2^=0.73, 95% CI 0.63–0.84). Participants read his arrival ‘from who knows where’ and his first words (‘You need a purpose, a plan, an idea!’) as the stance of a professional who imposes solutions without understanding the context. A psychologist wrote: ‘The Perfect One represents everything that, in my humble opinion, should not characterise a responsive carer’, adding that the responsive carer should be ‘that professional figure who listens, guides, informs, and makes parents and children aware and involved, truly making them protagonists of their own story’. Another noted that the Perfect One looks at first like what a mentor ‘should be’, and ‘from the very first description he reveals himself to be quite the opposite’.

Each valued misfit was associated with a distinct quality. The Punctured One scored highest on Emotional Processing (mean 1.31, SD 0.79), his holes read as the capacity to let difficult emotions pass: ‘Anger just passes through me: this represents the ability to let emotions flow, to not hold onto negativity.’ The Folded One and the Upside-Down One scored highest on Perspective Taking (means 1.33 and 1.92); participants read the Folded One’s creases as the capacity to hold each family’s history (‘He keeps all memories in his folds… ready to open them at the right moment, discovering unique treasures’), and the Upside-Down One’s view of ‘things others don’t see’ as a creative approach to families’ needs. The Wrong One scored high on Emotional Processing (mean 1.13, SD 0.74), and his rule that ‘when I manage to do something right, it’s time to celebrate’ was valued as a model of realistic expectations. Among the four valued misfits, scores differed on Emotional Processing (χ^2^=27.36, p<0.001; ε^2^=0.51, 95% CI 0.31–0.75), Perspective Taking (χ^2^=37.13, p<0.001; ε^2^=0.69, 95% CI 0.57– 0.83) and, to a lesser extent, Initiative/Activation (χ^2^=8.31, p=0.040; ε^2^=0.15, 95% CI 0.05–0.33).

### 3.5 The Feeble One

The Feeble One combined rejection with the highest Self-Reflection score of all characters (mean 1.00, SD 0.72) and the lowest Initiative/Activation score (mean −1.71, SD 0.46), on the dimension where the characters differed most (Kruskal–Wallis χ^2^=105.90, p<0.001; ε^2^=0.85, 95% CI 0.77–0.95). Participants found his passivity, ‘always tired, always asleep’, hard to reconcile with their norms of active engagement:

The misfit that repels me is the Feeble One. In contrast to the others, he doesn’t express himself (he collapses asleep). He evokes the image of a resigned carer, lacking enthusiasm, who settles into the context without wanting to make a difference.

The one I find hardest to tune into is the Feeble One, always tired, without objectives.

In the facilitated session 23 participants gave 59 emotional attributions (2.6 per participant), most often within the families of sadness (33.9%) and anger (28.8%), followed by fear and disgust (15.3% each), while only 3.4% fell within positive categories such as trust and acceptance. Anger and disapproval received the highest intensity ratings (median 7.5 each), followed by vigilance (median 7.0); irritation, the emotion mentioned most often in the chat (13 times, nine with a rating), had a median of 6.0 (range 5–7), and sadness the lowest median (5.0) with the widest range (5–9), so that for some participants the Feeble One evoked grief and for others a more contained reaction (Figure 3). Participants often reported several emotions at once, and some described a sequence:

**Figure 3.**
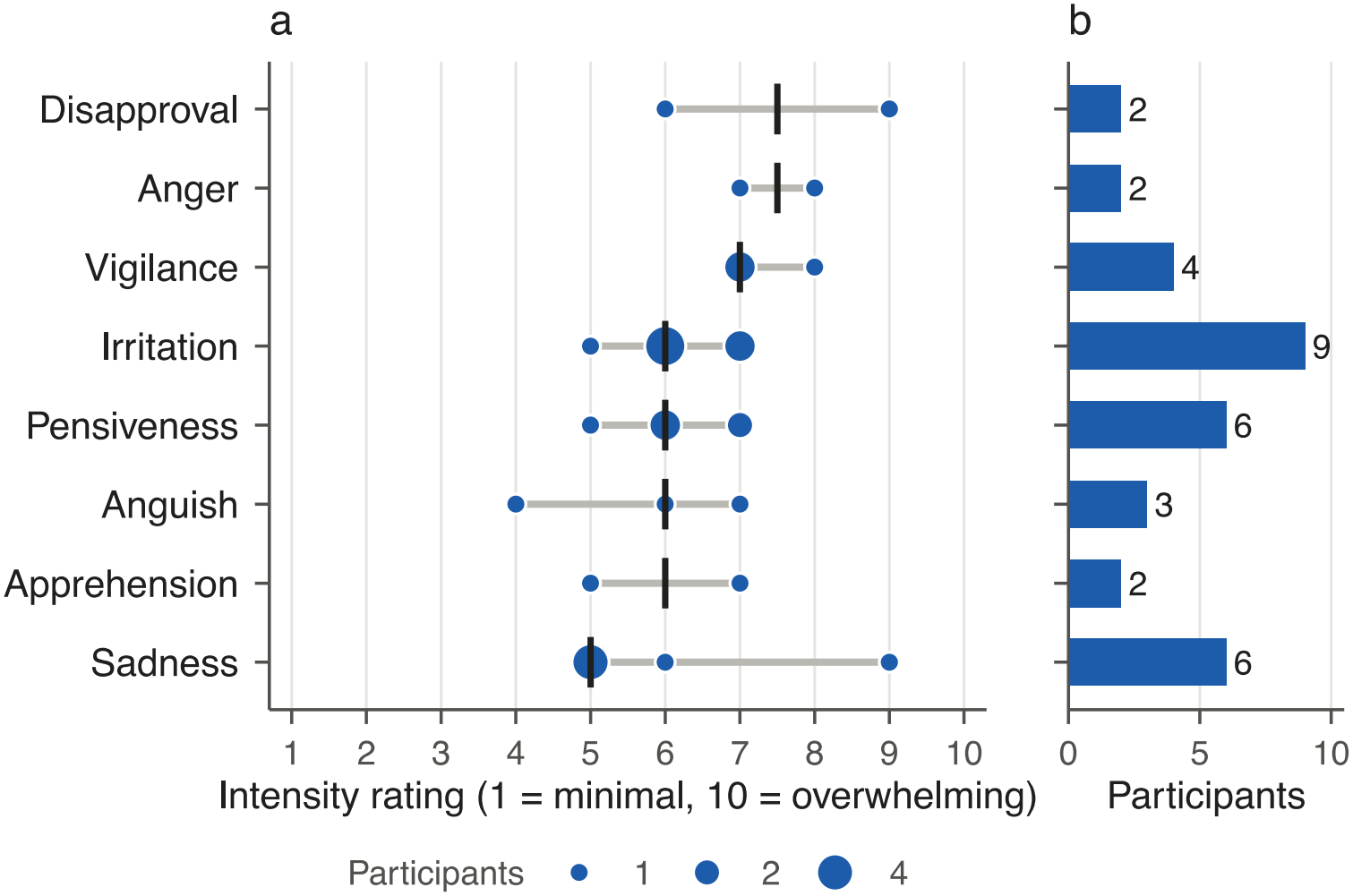
Emotional responses to the Feeble One during the facilitated Plutchik exercise (n=23 participants). (a) Intensity ratings: grey lines span the range of ratings, points mark the ratings given (point size proportional to the number of participants) and vertical bars mark the median. (b) Number of participants who reported each emotion with an intensity rating. Emotions are ordered by median intensity; only emotions rated by at least two participants are shown.

Irritation 6, vigilance 8, disgust 3, pensiveness 5, sadness 5

Disapproval 9 (alas, I’m being judgmental…)

At first it makes me angry 8, then a lot of anguish 7, and sadness 5

I think the Feeble One has so much insecurity (sadness 9)… if then I see that even working on it he stays feeble, then irritation 7 and anger 7

During the discussion one participant moved from ‘disapproval 9’ to ‘anguish 4, trust 7’, commenting ‘I hadn’t thought of the Feeble One in these terms’. Another wrote: ‘Looking at the situation from another angle that goes beyond the first reaction, I also feel sadness, pensiveness, apprehension.’ A third proposed to read the feeble caregiver as ‘welcoming, capable of accepting the other … non-judgmental, capable of adapting to the personality of whoever is in front of them’. The facilitator closed by distinguishing feebleness as a permanent state from a temporary condition: ‘Sometimes our feeble patient won’t remain feeble forever but needs time to function that cannot be that of the Perfect One… the time of healing, especially for mental illnesses, is not the time of the healthy.’

## 4 Discussion

### 4.1 Where the book evaluates: the Perfect One and the valued misfits

Participants adopted the verdicts that the book gives, often in its own words: the narrative dismisses the Perfect One as ‘a truly perfect fool’ and lets four misfits answer his contempt by naming their own strength, and participants rejected the first and valued the others. The rejection of the Perfect One was the most consistent finding of the study, across all dimensions and across professional groups. Participants identified him with a professional who positions himself as the sole bearer of knowledge and denies families recognition as experts in their own experience. Such a professional lacks what narrative medicine calls narrative competence, the capacity to acknowledge, interpret and act upon others’ stories (Gowda et al. 2019). One nurse observed that professionals ‘often position themselves as knowledgeable, and the patient frequently perceives themselves as inferior’, and noted that even the etymology of the word patient speaks of endurance and asymmetry; philosophical accounts of epistemic injustice describe the same hierarchies in medical language (Carel and Kidd 2014). Participants objected to expertise used in ways that silence families, and they described a responsive approach in which professional knowledge is held in relation with families and supports them without directing them (Ravaldi et al. 2024). Several read the Perfect One’s arrival ‘from who knows where’ and his final isolation as a warning about expertise that belongs to no relational context. Over recent decades clinicians and health systems have moved from paternalism towards shared decision-making (Mulley et al. 2012), and in difficult perinatal conditions shared decisions are associated with higher maternal satisfaction (Ravaldi et al. 2023b). We read the unanimity of the rejection as evidence that paternalism has lost normative legitimacy in this cohort, and we do not take it as evidence that paternalism has left everyday practice, where its subtler forms may persist.

Participants assigned the qualities of responsive care to different valued misfits, and none of the misfits held all of them. We call this pattern distributed competencies: participants placed the relational capacities of responsive care across persons, contexts and moments, and several said so in terms of teamwork. A midwife wrote that it is ‘absolutely necessary to work in multi-disciplinarity… to place ourselves as a system, where there are many parts, equally important, that only together can achieve the intended goal’. Another wrote: ‘No leader can be such without a winning team. We must remember that no one is perfect and omniscient.’ Studies of interprofessional collaboration likewise tie good outcomes to teams whose members contribute complementary strengths (Reuter et al. 2018; Zaher et al. 2022). If responsive care is distributed among people, the idea of the ideal caregiver has to change. Traditional models of professional excellence imagine one practitioner who holds every competency to a high degree; in Alemagna’s fable the character who claims that completeness ends alone, and participants read his fate as a warning. In professional education, a model of the ideal professional who holds every competency places on practitioners a demand that the Perfect One’s isolation illustrates, and practitioners who cannot meet every need alone may read the gap as a personal failure. In the misfits’ model, each practitioner contributes a characteristic strength and depends on colleagues for the others, a pattern that may also be easier to sustain over a working life. One participant saw it in the Punctured One, who ‘is responsive because he sees the difficulties of all of them and responds by stimulating each to find their best personal solution, without being judgemental’. In this view practitioners become competent by accepting their own imperfection and integrating it within a web of complementary relationships.

We read these results as evidence about the book as much as about the participants, because where Alemagna’s characters carry an explicit evaluation, participants’ judgements largely followed it, and in the forum they voiced norms of care that the author had already stated.

### 4.2 Where the book is silent: the Feeble One

The Feeble One is the only misfit who claims no virtue, since he falls asleep when the misfits are asked what they can offer and leaves with the others without his place in the group being explained. With this character participants’ reactions departed most clearly from the book’s direction, and since the narrative offered no evaluation, their judgements drew on their own professional norms of activity and responsiveness. One participant wrote: ‘The character I find irritating is the Feeble One, who can sleep even while the Perfect One criticises him harshly. I am repelled by his passivity and complete lack of response to stimuli. Perhaps he repels me because this attitude lurks deep within me, and I often fear it might take over.’ The qualities participants found most aversive were the ones they recognised in themselves.

We describe this reaction as projective disavowal, the simultaneous recognition and repudiation of one’s own vulnerability to passivity, exhaustion and withdrawal. A midwife linked it to the culture of her work: ‘we live in a performative culture where “not doing” or even just “not managing to do everything” is often seen as something feeble and weak. Perhaps I should be more welcoming towards the Feeble One, who makes it legitimate to take breaks.’ Participants also separated the Feeble One as colleague from the Feeble One as patient. A nurse in neonatal intensive care said: ‘If I think of him as a caregiver, I could never work with him; I already struggle with patients… but they are patients!’ Participants tolerated passivity in patients and found it intolerable in colleagues, and in doing so they stated an expectation of constant activity that they also applied to themselves. Practitioners who work under norms of constant productivity may feel no permission to acknowledge their own depletion, and they may bring the same impatience to withdrawn or depressed parents.

### 4.3 Facilitated discussion of reactions to the Feeble One

We did not use *The Five Misfits* as a text about healthcare, since picture books of this kind offer symbolic figures and provoke affective responses without prescribing professional action (Ravaldi and Vannacci 2025), and readers negotiate emotional and professional meanings in them much as in the literary texts of narrative medicine, and the facilitator used this distance in a session that moved through four phases.

In the first phase the facilitator legitimised aversive emotions, presenting Plutchik’s wheel as a description of the whole human range of emotions (‘Plutchik started from fundamental emotions and then graduated them, combining them into different constructs’), so that the discussion moved from moral judgement to observation and participants could name irritation, disgust and vigilance without defending themselves (‘Grading one’s own emotions is a beautiful work of awareness: clearly, there are no emotions more beautiful than others’). We see this step as close to Charon’s account of narrative medicine, in which clinicians attend to their own responses to patients’ stories (Charon 2001). She then reframed the reactions clinically, observing that ‘the risk is not being able to work at the maximum of our competencies because of this wave of emotions we feel with women or fathers with depression’. Participants began to describe parents with masked depression, emotional withdrawal or apparent disengagement. One observed that ‘the Feeble One can be imagined as a carer, or as a patient, or simply as a human being’, and the discussion turned to who is allowed to be feeble in healthcare settings. Some questioned the expectation that professionals should stay unaffected: ‘There’s the problem of considering the caregiver as someone who should be non-human and not have bad moments.’ The group came to treat professional vulnerability as part of the work, shared by everyone in it.

In the third phase participants distinguished forms of feebleness that look alike and call for different responses. A psychologist observed that ‘behind feebleness there may be aggression that demands constant care, or it may be an expression of major depression, and the emotions they evoke are different’. In perinatal mental health, passivity and withdrawal may signal exhaustion, depression, hostility or dissociation, and Norman et al. (2007) count contextual discrimination of this kind among the marks of clinical expertise. Participants extended the distinction to institutions, describing organisational inertia and outdated systems as ‘feeble’ and draining, and the facilitator acknowledged that practitioners deal with feeble institutions as well as with difficult patients; one participant summed it up as ‘there exists feeble within feeble’. Fiction reached this competence by a route of its own: participants first reacted strongly to a character and were then guided to examine and differentiate their reactions. In the last phase participants reworked their first reactions: some revised their ratings in public (‘I’m changing from disapproval to anguish and trust’); one asked whether vigilance might also mean ‘monitoring and careful attention’, which led the group to discuss where protective attunement ends and overcontrol begins; others read feebleness as receptivity, and with it questioned professional cultures that equate value with constant activation. The fictional character gave them a surface on which to project reactions that would be hard to state about real patients or colleagues, and the facilitator’s guidance turned those reactions into distinctions they could use in practice.

### 4.4 Limitations

The study involved a single cohort of women attending one course, and participants chose which misfits to discuss, so that the number of reflections varies across characters. The scoring was done by the research team after an AI-assisted first pass, and the two composite indices of the map are descriptive constructions. We did not measure outcomes over time, and we cannot say whether the changes of view observed in the session lasted.

## 5 Conclusions

Facilitated reading of *The Five Misfits* brought tacit professional attitudes of perinatal practitioners into view. Participants rejected the Perfect One as an anti-model of care and described responsive practice as a set of competencies distributed across persons and contexts, in both cases following the evaluations the book itself provides. The Feeble One, whose value the book leaves unstated, drew intense aversion together with the highest self-reflection, and through him participants confronted difficult patients, difficult colleagues and their own vulnerability to exhaustion and withdrawal. In the facilitated session they named these reactions, related them to parental depression and learned to distinguish forms of withdrawal that call for different responses. The session moved through four phases, from the legitimisation of aversive emotions through clinical reframing and differentiated understanding to an integrative revision of first reactions, and the facilitator guided each passage from one phase to the next. We call the aim of this work sustainable professional presence: the capacity to remain emotionally available to families without depleting one’s own resources.

Picture books let readers meet emotionally charged material at a safe symbolic distance, where implicit professional reactions can surface and a group can examine them together. For arts-based education in health, we suggest that those who choose a work should weigh its silences as well as its messages. When a book evaluates its characters, readers tend to agree with it, as our participants did with the Perfect One and the four valued misfits. When the author leaves a character unexplained, readers fall back on their own norms, and a skilled facilitator can make those norms the subject of reflection. Programmes for perinatal professionals should include such exercises, with time for facilitated discussion, and further studies should test whether they sustain professionals’ presence with withdrawn or depressed parents and protect against burnout.

## Data Availability

Data are available upon reasonable request.

## Acknowledgements

We wish to express our gratitude to all the participants of Florence University “*I Primi Mille Giorni*” course 2024-25, who contributed to the forum discussions and participated in the in-class dialogues that inspired the reflections elaborated in this work. While their contributions do not qualify them for authorship, we deeply appreciate their engagement with the themes presented in this study: Arianna Balestracci, Giada Barbirato, Elena Baritello, Simona Bensi, Elisa Bertilorenzi, Francesca Casartelli, Chiara Castracani, Marina Ceragioli, Cecilia Cirilli, Valentina D’Alesio, Giulia Fava, Anisia Fazzi, Irene Graziani, Marcella Marcone Terzago, Nausicaa Martino, Chiara Mauri, Federica Minelli, Albina Mingaj, Valentina Modolo, Tosca Mori, Miriam Morici, Laura Mosconi, Alice Pasquini, Noemi Passalacqua, Chiara Poli, Anna Pomes, Chiara Rossini, Althea Iris Salerno, Valentina Scarpello, Ambra Sciascia, Maria Selene Tarascio, Silvia Zanette, Alessandra Zani, and Laura Zanoni.

## Declarations

### Author contributions

Claudia Ravaldi: conceptualisation, investigation, writing (original draft, review and editing), project administration. Alfredo Vannacci: conceptualisation, investigation, data curation, formal analysis, writing (original draft, review and editing).

### Declaration of generative AI use

ChatGPT 5.2 (OpenAI) and Claude Sonnet and Opus 4.5 (Anthropic) supported the initial coding and the thematic quantification of the forum discussions, as described in the Methods, and the translation of specific passages from Italian into English; Claude Opus 5.5 (Anthropic) supported the editing and shortening of the manuscript. The authors directed and checked every use of these tools and take full responsibility for the content of the article.

### Data availability

The data that support the findings of this study are available on request from the corresponding author. The data are not publicly available due to privacy restrictions, as they consist of participants’ written reflections.

